# A Natural Language Processing Pipeline to Study Disparities in Cannabis Use and Documentation Among Children and Young Adults A Survey of 21 Years of Electronic Health Records

**DOI:** 10.1101/2022.10.12.22281003

**Authors:** Nazgol Tavabi, Marium Raza, Mallika Singh, Shahriar Golchin, Harsev Singh, Grant D. Hogue, Ata M. Kiapour

## Abstract

The legalizations of medical and recreational cannabis have generated a great deal of interest in studying the health impacts of cannabis products. Despite increases in cannabis use, its documentation during clinical visits is not yet mainstream. This lack of information hampers efforts to study cannabis effects on health outcomes. A clear and in-depth understanding of current trends in cannabis use documentation is necessary to develop proper guidelines to screen and document cannabis use. Here we have developed and used a hierarchical natural language processing pipeline (AUROC=0.94) to evaluate the trends and disparities in cannabis documentation on more than 23 million notes from a large cohort of 370,087 patients seen in a high-volume multi-site pediatric and young adult clinic over a period of 21 years. Our findings show a very low but growing rate of cannabis use documentation (<2%) in electronic health records with significant demographic and socioeconomic disparities in both documentation and use, which requires further attention.

## Introduction

In the United States, cannabis is legal for medicinal use in 37 states, and legal for recreational use in 19 states. Up to 22-million Americans twelve years old or older use cannabis annually. This is part of an upwards trend. Daily reported usage of cannabis is increasing, from 2.1% in 2016 to 3.4% in 2019 according to the national survey on drug use. Within this population, there has also been increased usage among youth [1]. Over 11.8 million young adults report cannabis use. Daily use has increased from 5.9% to 6.9% for twelfth graders, 2.9 to 4.4% for tenth graders, and 0.8 to 1.1% for eighth graders from 2017 to 2020 [2].

Despite this increase in usage, proper documentation of cannabis use has not become mainstream. Such information is vital for a more accurate assessment of cannabis use rates and potential effects on treatment outcomes. For example, cannabis has shown to influence remodeling of a range of musculoskeletal tissues (e.g., bone) [3]. A recent study on pediatric patients with extremity fractures found significantly increased time to union in those who used cannabis [4]. There is also evidence suggesting cannabis use affects the outcomes of surgeries, including mortality, pain, comorbidities, and revision rates [5, 6]. One study found reduced mortality in cannabis users undergoing total hip arthroplasty, total knee arthroplasty, total shoulder arthroplasty, and traumatic femur fixation [7]. Other studies found that cannabis users had higher surgery revision rates [8] and cannabis users undergoing spine surgery had greater perioperative morbidity [9].

In terms of pain after surgery, results are contradictory as well. Cannabis users reported lower pain in the operative site in one study of 937 patients [10], while patients with preoperative cannabis use reported increased pain after major orthopedic surgery on an NRS scale in another study of 3793 patients [11]. A different study found that cannabis users had higher total prescribed opioids, and longer duration of use [12]. A recent review found that cannabis use in the form of combustive cigarettes represents perioperative risks for induction/anesthesia, post operative pain, and analgesia in teenagers [13].

Currently the most common way of cannabis use documentation in patients’ health records is within unstructured clinical notes, in an unstandardized manner [14]. While studies have proposed the development of standardized screening protocols to streamline documentation [15], yet no such protocol has been adopted widely. Currently, cannabis itself is referred to in heterogeneous ways within clinical notes, alternatively referred to as MJ, cannabis, weed, CBD and THC. Some of these terms, specifically weed and CBD, are nonspecific (i.e., weed also refers to other plants, CBD also refers to common bile duct). These terms may also be misspelled. In addition, particularly within pediatric clinical notes, there are mixed mentions of cannabis use among patients and their family members. Additional analysis is needed to distinguish which subject is the cannabis user. Overall, the complexity of retrieving cannabis use screening information from this type of heterogeneous, unstructured data has made it difficult to monitor or study.

One approach to tackle this challenge is the use of natural language processing (NLP) to identify cannabis users. NLP uses linguistic knowledge to identify information (such as cannabis use) from human language input by identifying patterns in the data. Applying NLP techniques on clinical notes comes with its own challenges such as presence of noise, heterogeneity, different templates in the data, abbreviations, misspellings, incomplete sentences, etc. Hence, many of the state-of-the-art NLP models do not perform as well when applied directly to clinical notes and the data needs to be thoroughly preprocessed and cleaned before being fed into the models. Studies such as Tavabi et al. [16], Wang et al. [17], Ling et al [18], developed and evaluated such NLP pipelines and approaches on clinical notes for purposes like cohort identification and building registries.

Some studies have also used NLP to identify substance use from clinical notes. One study used NLP on clinical notes to identify hospitalized trauma patients with alcohol misuse and demonstrated greater accuracy than EMR-based billing codes [19]. Another study detected alcohol, drug, or nicotine use from unstructured notes using NLP and achieved good performance over a wide breadth of notes [20]. When looking at cannabis use specifically, a separate study developed an NLP algorithm to identify cannabis related terms, historical mentions, and hypothetical mentions within EHR notes [21]. However, Carrell et al.[22] identified 54% of the notes with positive medical cannabis usage automatically and used an NLP-assisted manual review tool to identify the rest, which is a labor and time intensive process.

In this work, we developed an NLP pipeline to comprehensively assess the changes in cannabis use documentation over the past 21 years along with potential disparities in use and documentation in a multi-site high volume pediatric and young-adults orthopedic and sports medicine (OSM) academic practice, without the need for manual chart review. We looked at patients from the OSM considering the high prevalence of these injuries among children and young adults, and effects of cannabis in musculoskeletal tissue healing and remodeling. With a better understanding of how cannabis use is documented, care guidelines for a growing population of cannabis users can improve.

## Methods

This study was performed in line with the principles of the Declaration of Helsinki. Approval was granted by the Institutional Review Board of Boston Children’s Hospital. Considering the retrospective use of available data, the study was exempt from patient consent. A breakdown of the pipeline developed in this study is shown in Figure 1. First, through a process of physician-in-the-loop a dictionary of cannabis related keywords and their possible misspellings was generated. Based on the dictionary, a dataset of clinical notes was extracted, and after preprocessing and filtering out irrelevant notes, the sentences with cannabis related information were retrieved from the notes. A small percentage of the sentences were then annotated by a group of experts to generate training data for a classifier to confirm positive vs negative cannabis use. Afterwards, the structured Electronic Health Records (EHR) of the case and control group (patients with predicted positive and negative cannabis usage) such as demographics and list of diagnoses and procedures were extracted and used for analysis.

**Figure 1:**
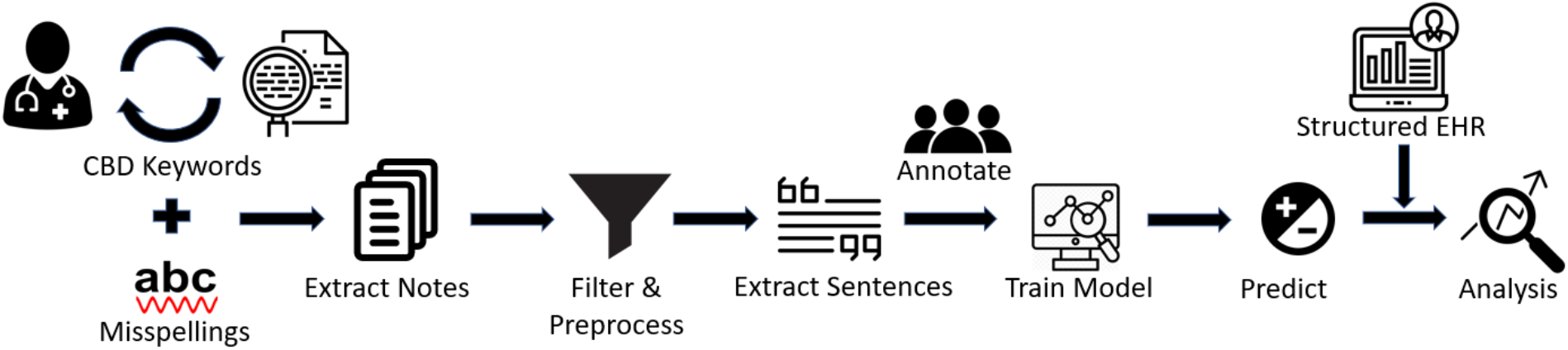
Diagram of pipeline developed in this study to evaluate cannabis use documentation and to assess disparities in cannabis use among children and young adults with musculoskeletal injuries.

### Data

Following IRB approval, all clinical notes, baseline demographics, and diagnostics and procedures of every patient who has visited the Boston Children’s Hospital OSM clinics (6 locations across Massachusetts) between 2000-2021 were obtained (370,087 unique patients, 23,871,108 notes). The baseline demographics of the cohort are presented in Table 1. The Social Vulnerability Index (SVI) was calculated based on residence zip code. SVI is a composite index developed by the Centers for Disease Control and Prevention that characterizes community resilience and vulnerability relative to external stressors [23]. It calculates an overall index (0 – 1), with a higher index indicating greater social vulnerability. Additionally, current procedural terminology (CPT) billing codes were used to identify most common musculoskeletal procedures within the study population and diagnosis codes were used to identify patients with cannabis related diagnosis.

**Table 1.**
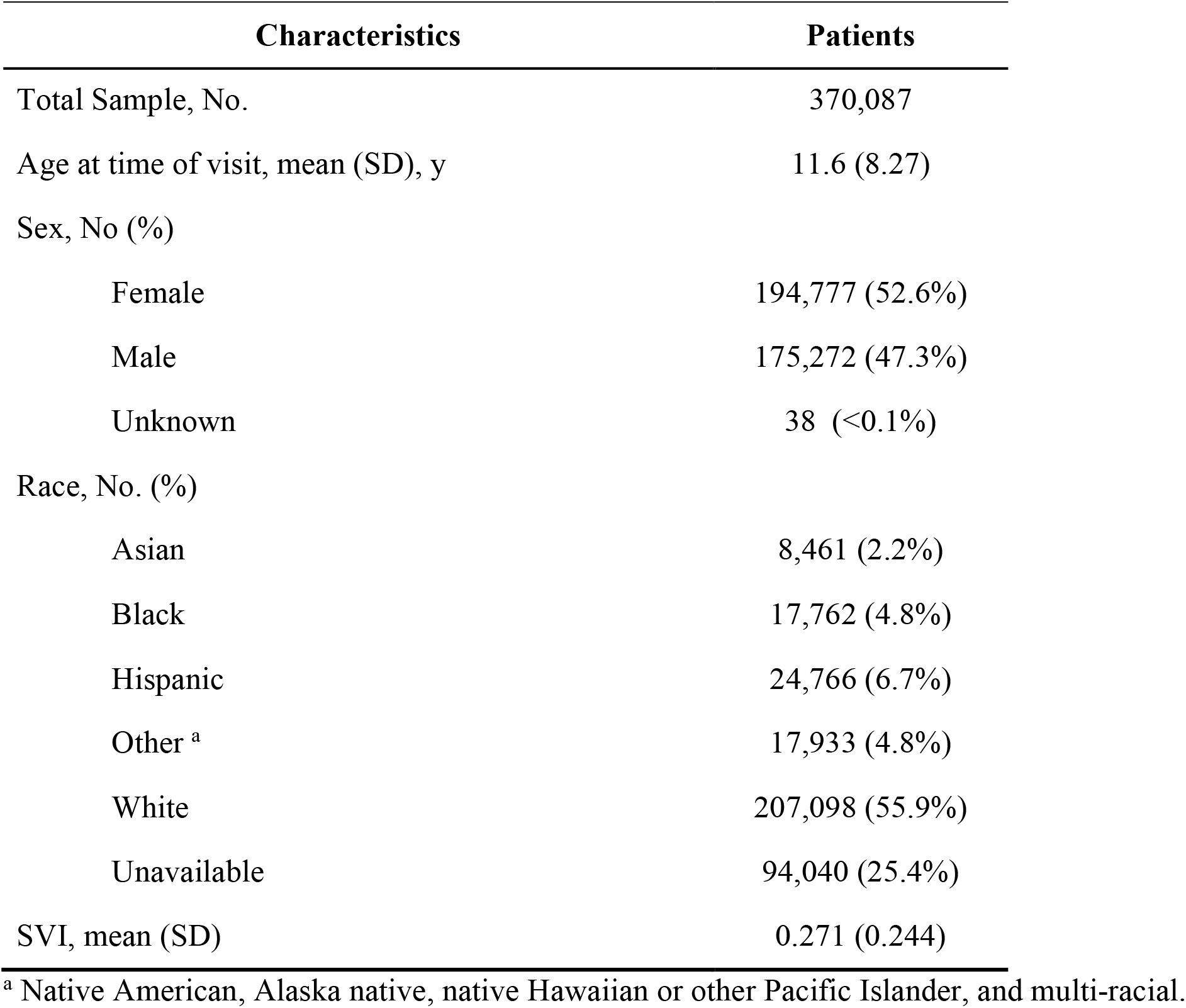
Baseline characteristics of the studied cohort.

### Diagnostic Codes for Cannabis Misuse

ICD (International Classification of Diseases) and SNOMED CT (Systematized Nomenclature of Medicine Clinical Terms) are 2 different coding systems used for diagnosing patients. ICD codes are mostly used after the care is completed for example for billing purposes, whereas SNOMED CT is used directly by healthcare providers during the process of care. There are different ICD codes and SNOMED CTs related to cannabis use. To identify them, we extracted all diagnoses with the keyword “cannabis” in their description, and after they were reviewed and confirmed, all cases with those codes and all the patients diagnosed with them were extracted. Table 2 shows identified diagnostic codes, their descriptions, and their frequency. A patient might get diagnosed with the same code or a similar code more than once.

**Table 2.** Diagnostic codes related to cannabis use found in the dataset. Diagnostic codes with frequency less than 10 are not shown in the table.

| Diagnosis Code | Source Vocabulary | Description | Count |
| --- | --- | --- | --- |
| F12 | ICD-10-CM | Cannabis related disorders | 10543 |
| 305.2 | ICD-9-CM | Nondependent cannabis abuse | 6990 |
| 304.3 | ICD-9-CM | Cannabis dependence | 4832 |
| T40.7 | ICD-10-CM | Poisoning by, adverse effect of and underdosing of cannabis (derivatives) | 142 |
| 37344009 | SNOMED CT | Cannabis abuse | 136 |
| 428823006 | SNOMED CT | Cannabis misuse | 65 |
| 85005007 | SNOMED CT | Cannabis dependence | 53 |
| 191837001 | SNOMED CT | Cannabis dependence, continuous | 12 |

In the ICD coding system, there is a hierarchical structure. By adding more characters to the code, the diagnosis becomes more specific. For example, for F12 in Table 2 the actual diagnosis codes were F12.20, F12.10, F12.90, etc. Here they are aggregated to the most general cases (e.g., parent codes) related to cannabis use.

### Natural Language Processing (NLP) Model

A hierarchical NLP approach was developed to first identify notes containing cannabis-related terms (screening step) and then classify them into positive (endorsed cannabis use) or negative (no cannabis use).

For the screening step, first a cannabis-related dictionary was generated with a physician-in-the-loop approach. Initially A few seed words (Marijuana, Cannabis, CBD, Weed, THC) and medical terms (Tetrahydrocannabinol, Epidiolex, Cannabidiol, …) from FDA approved cannabis drugs were chosen and notes containing them were collected. Afterwards, based on the notes collected, other keywords (such as MJ) were identified and added to the dictionary.

Additionally, possible misspellings of the keywords were identified and added to the dictionary. In order to add misspellings of dictionary keywords, words from all the notes were gathered and their N-grams (N=[1-3]) were computed. The cosine similarity of each word’s Ngram and Ngram of the correct word was computed, and words with higher similarity than certain threshold were identified. The threshold was chosen manually for each keyword in the cannabis-related dictionary. Below is an example of how this is computed.

*A = Marijuana N-grams (A:3, JU:1, AH:0…)*

*B = Marijauna N-grams (JA:1, JU:0, AU:1, JAU:1…)*

*C = Marijuahana N-grams (A:4, H:1, AH:1, AHA:1…)*

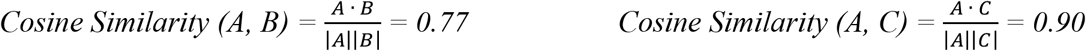

When the dictionary was finalized, all the notes in the dataset (n=23,871,108) were screened for cannabis-related terms.. Figure 2 shows the number of notes identified with each keyword (some notes contained more than one keyword).

**Figure 2.**
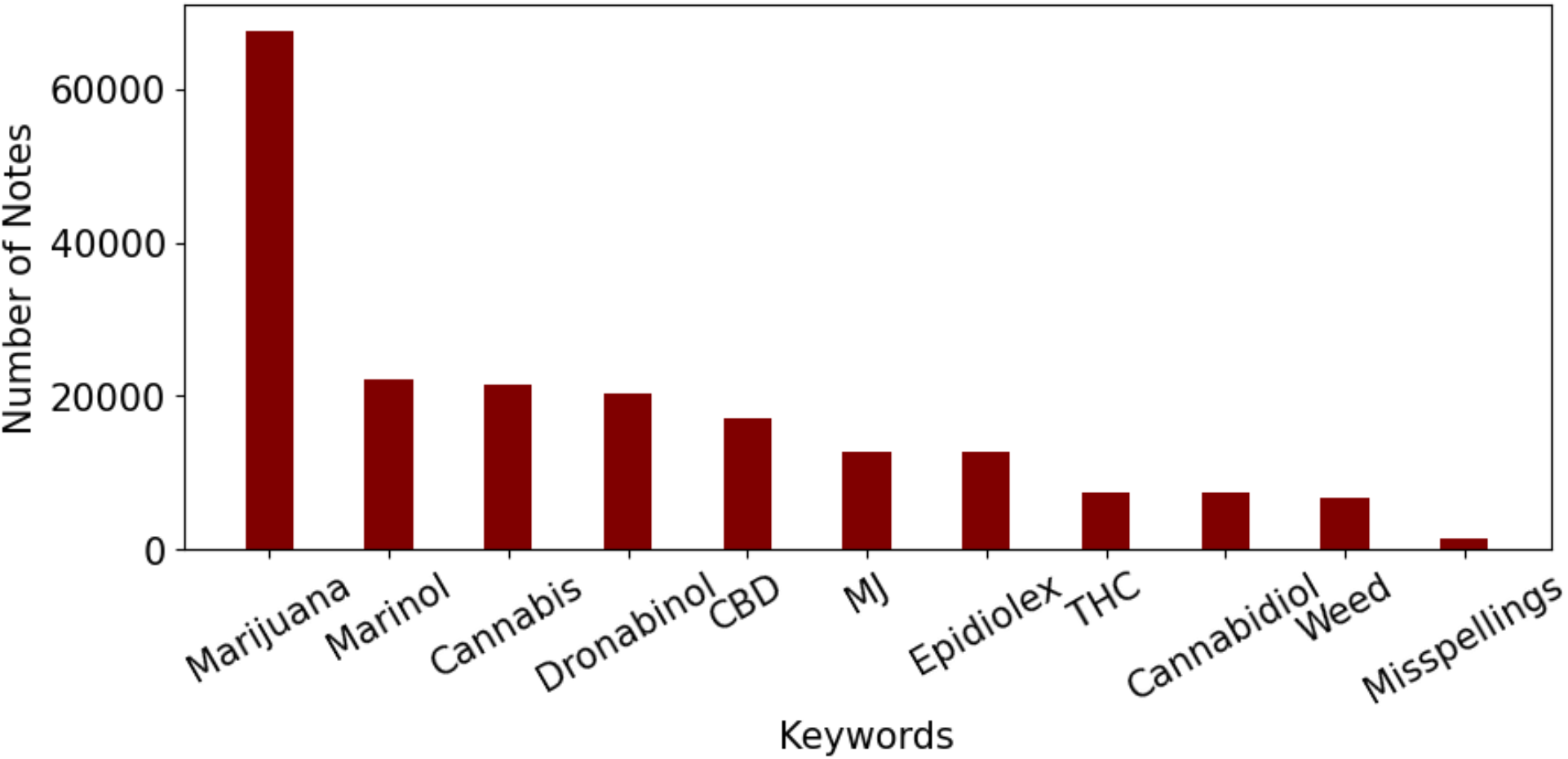
The keywords used to extract clinical notes and the number of notes containing each of them. Keywords like Syndros and Cesamet which appeared in less than 10 notes are not included in the graph.

After finding the notes with any of the keywords in the dictionary, the dataset was first cleaned and processed. The first step was to remove the notes that were incorrectly marked as cannabis related. As mentioned earlier words like CBD or Weed can also be used in other contexts. The acronym CBD is also used for Common Bile Duct and is used in notes related to gastrointestinal conditions. Also the keyword Weed is used in notes related to allergy detection. To remove these irrelevant notes, 2 other sets of keywords were compiled. Words like abdominal, gallbladder, pancreas, common bile duct, … were used to identify and filter out notes related to common bile duct and words like pollen and allergy and its variations were used for allergy related notes. Additionally, cannabis related notes which correspond to very young patients (under 7) were primarily related to the cannabis use in their household and not by the patient, especially during pregnancy. Table 3 shows a few examples for patients under age 7. Since the purpose of this study is to identify patients with direct cannabis use, patients younger than 7 years old were removed from the dataset.

**Table 3.**
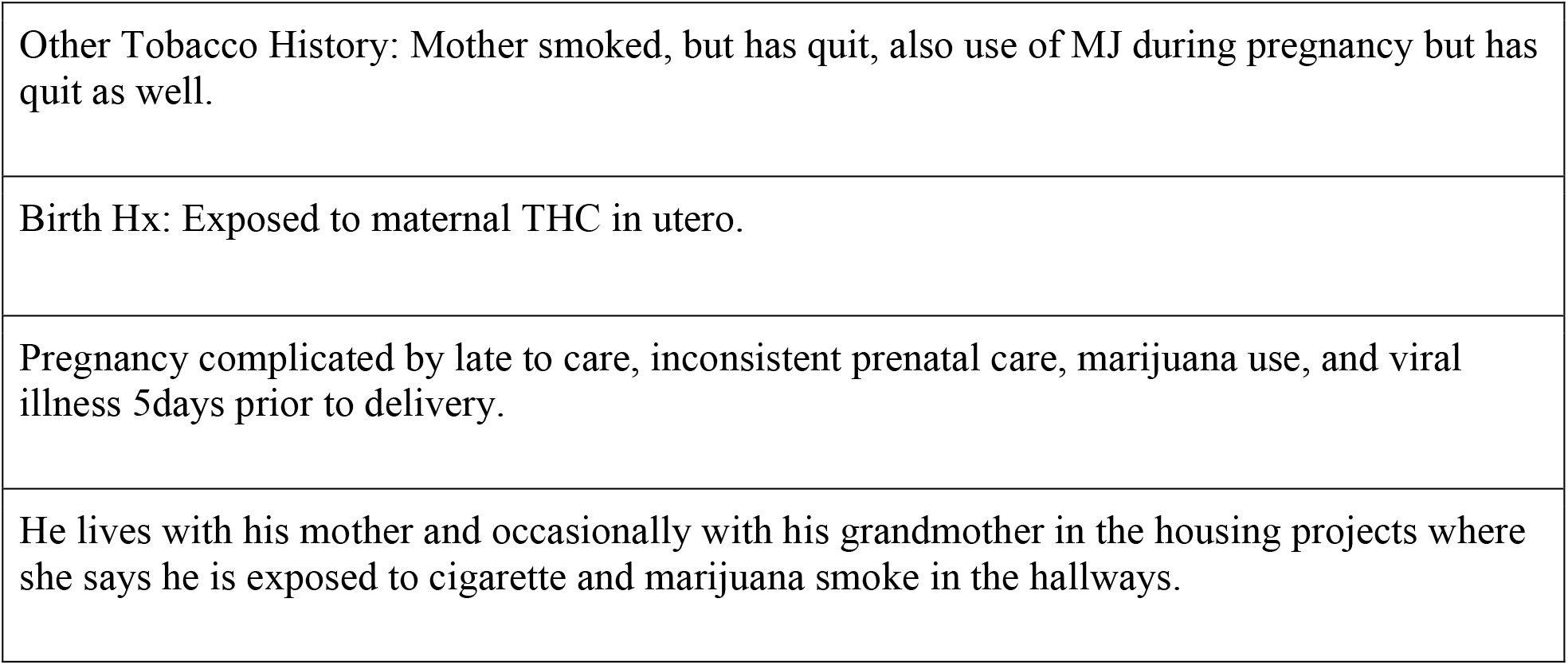
Examples of Cannabis Related Notes for Patients under 7.

After preprocessing the notes, the sentences containing keywords from the cannabis dictionary were extracted from each note. The types of extracted notes were very heterogeneous, resulting in a heterogeneous collection of note contents and formats. Some notes had forms template with different fields and without proper sentences and in some others, the note contained full sentences with a lot of detailed narratives. Since the cannabis use was only a small part of the whole notes and to minimize unwanted bias related to note types (i.e., template and content), only the sentences containing the keywords were extracted and used as input to the classifier.

After the screening step, the model Bidirectional Encoder Representations from Transformers (BERT) model was used to classify the extracted sentences. BERT is the state-of-the-art NLP model developed and trained by Google, which has been widely used across multiple domains, including healthcare [23]. BERT models provide contextual representations of words and sentences, which can then be used in classification. Domain specific BERT models yield significant improvement, hence for this project we used ClinicalBioBERT [24], which has been pretrained on medical literature (pubmed corpus) and publicly available MIMIC dataset [25]. We further pretrained the ClinicalBioBERT on all the available notes from Boston Children’s Hospital (BCH_BERT; 23,871,108 notes for 1 epoch) so the model would be more familiar with the language used in the clinical notes. We then used the following deep learning model (Figure 3) to classify the extracted sentences from the screening step into positive use (i.e., self-reported cannabis use by the patient or guardians currently or in the past, positive use reports by the clinical care team or toxicology reports) or negative use (i.e., patient or guardian reported no use, negative use confirmation by clinical care team or toxicology reports, discussions of pros and cons of cannabis use with no direct indication of positive use, reported abuse for someone other than the patient). Examples of different types of both positive and negative use are given in Table 4.

**Table 4.** Example notes of positive and negative cannabis use. The green color shows positive cannabis cases and the red color shows negative.

|  |
| --- |
| Pt admits to smoking marijuana daily |
| MJ: + |
| agreed to try marinol with relief, continued scopolamine patch and all other antiemetics changed to prn. Be aware you may be light-headed or dizzy while using marinol RX: DRONABINOL (MARINOL( 2.5MG/TAB |
| Denies drugs or alcohol; states used marijuana "one year ago". Urine tox screen: positive for amphetamines and cannabis. |
| He has not tried any other drugs and denies any use of marijuana, cocaine, heroine, or other drug abuse. |
| Labs - Urine tox sent - negative for amphetamines, barbiturates, benzos, cannabis, cocaine, opiates, PCP Wrist films and R hand film done - Negative for fracture |
| I discussed the interactions of stimulant medications with alcohol, which increases the concentration of the stimulant and marijuana, which increases the heart rate with use of a stimulant medication. |
| CRAFFT score is 1; he has been in the car when his brother has been smoking marijuana and driving, but his brothers will not allow him to do this. |

**Figure 3.**
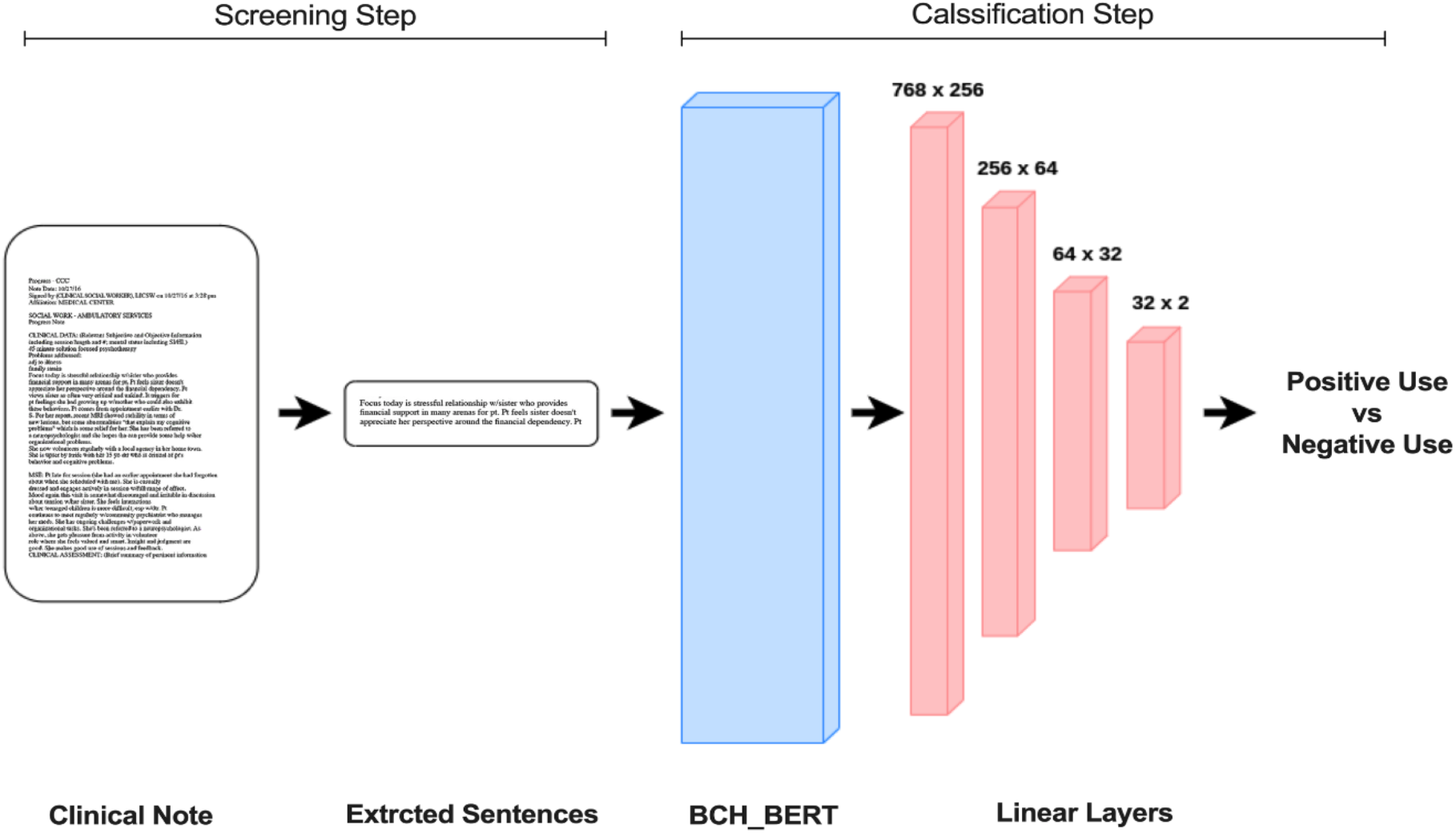
The deep learning model used to classify extracted sentences to identify positive cannabis use.

The model was trained and tested on a diverse set of manually labeled notes (n=3,835, 73% positive, 27% negative, 80% train, 10% validation, 10% test). The training was done for 7 epochs on an NVIDIA Titan RTX GPU. The model achieved an accuracy of 0.95, area under the ROC curve of 0.94, sensitivity of 0.97 and specificity of 0.90 on the test set.

### Evaluation of Disparities in Documented Cannabis Use

Disparities in the data were evaluated in 2 different settings. First, patients with documented cannabis use vs patients with no documents mentioning cannabis use. Second, patients with positive cannabis use vs patients with negative cannabis use. In the first setting the group with no documented cannabis use were never asked about cannabis usage and cannabis was not discussed in any of their notes whereas the other group had mentions of cannabis in at least one note which could be either a positive or a negative use.

A generalized linear mixed model was used to evaluate the associations between sex, race and SVI in rate of documented cannabis use (first setting). Adjusted odds ratios (aOR) were calculated and considered significant at P<.05 (SPSS v27) between those with documented cannabis use (positive or negative) and those with no cannabis use documentation. The aOR was calculated for female sex compared to male sex, and for asian, black, hispanic and other races compared to white race. For SVI, the aOR was calculated for every 0.01 point change in SVI. The analysis was repeated after excluding those with medical cannabis use. The same framework was applied to assess disparities in positive vs negative cannabis users with regards to sex, race and SVI (second setting).

## Results

Of 23,871,108 notes (7.8% OSM notes), 166,530 (2.7% OSM notes) contained cannabis-related terms (23,974 patients), out of which 124,952 notes (2.3% OSM notes) deemed positive use (13,556 patients). The breakdown of patients with positive and negative cannabis use are presented in Table 5. Of all identified positive patients, only 1,971 (14.5%) had medical diagnosis of cannabis use disorder based on either ICD codes or SNOMED CT (Table 2). From 2000-2021, there were increases in overall and positive cannabis use documentation across all notes (Figure 4). There were stepwise increases in cannabis use documentation across all notes after legalization of medical (2012, black vertical dotted line) and recreational (2016, green vertical dotted line) cannabis (Figure 4). For the OSM notes, there were increases in overall and positive cannabis use documentation primarily after 2010 with marked increases in cannabis use documentation after legalization of recreational cannabis (2016, green vertical dotted line; Figure 4).

**Table 5.**
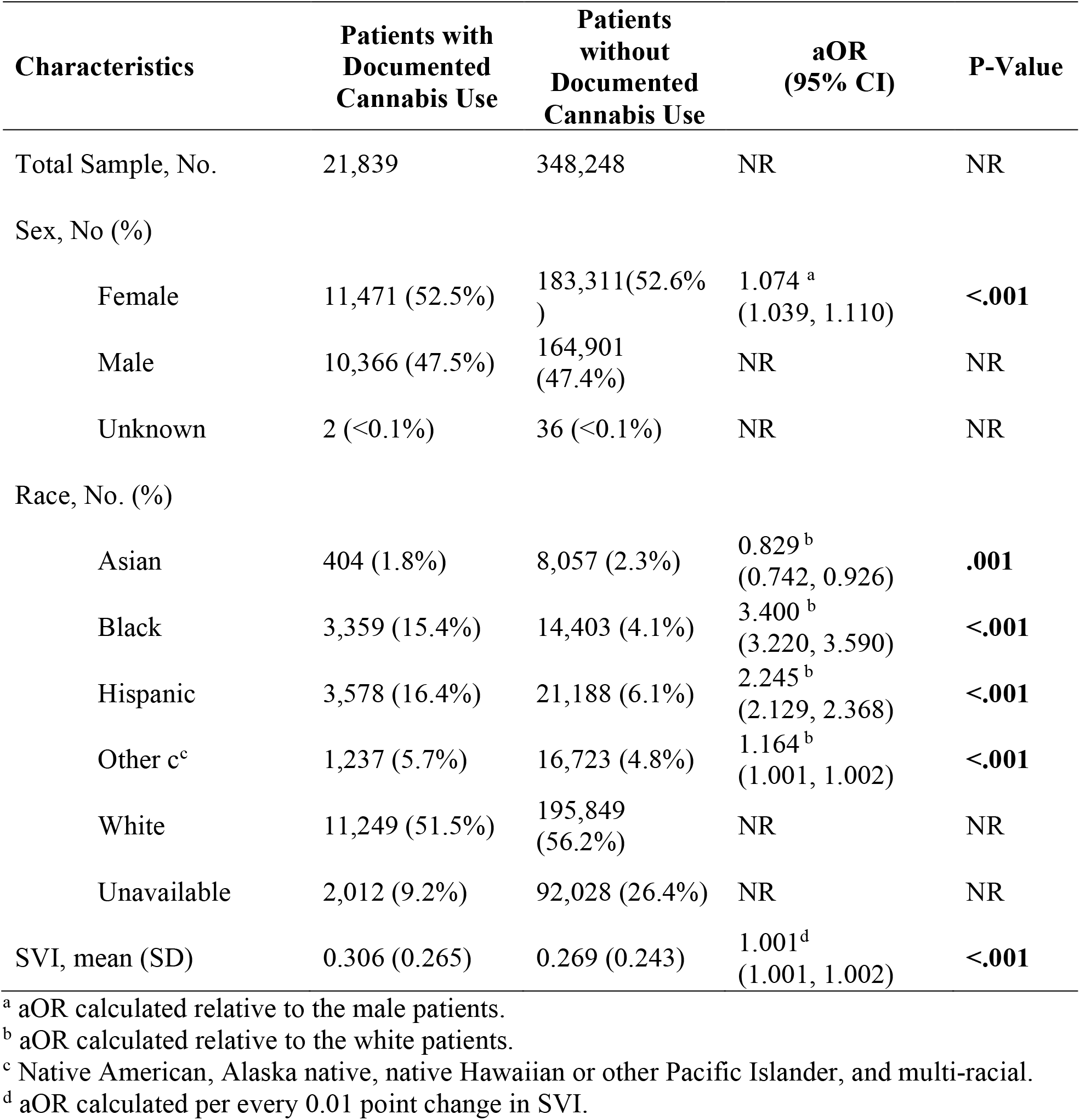
Characteristics of patients with and without cannabis use documentation.

| Characteristics | Patients with Documented Cannabis Use | Patients without Documented Cannabis Use | aOR (95% CI) | P-Value |
| --- | --- | --- | --- | --- |
| Total Sample, No. | 21,839 | 348,248 | NR | NR |
| Sex, No (%) |  |  |  |  |
| Female | 11,471 (52.5%) | 183,311(52.6%) | 1.074<sup>a</sup><br>(1.039, 1.110) | **<.001** |
| Male | 10,366 (47.5%) | 164,901 (47.4%) | NR | NR |
| Unknown | 2 (<0.1%) | 36 (<0.1%) | NR | NR |
| Race, No. (%) |  |  |  |  |
| Asian | 404 (1.8%) | 8,057 (2.3%) | 0.829<sup>b</sup><br>(0.742, 0.926) | **.001** |
| Black | 3,359 (15.4%) | 14,403 (4.1%) | 3.400<sup>b</sup><br>(3.220, 3.590) | **<.001** |
| Hispanic | 3,578 (16.4%) | 21,188 (6.1%) | 2.245<sup>b</sup><br>(2.129, 2.368) | **<.001** |
| Other c<sup>c</sup> | 1,237 (5.7%) | 16,723 (4.8%) | 1.164<sup>b</sup><br>(1.001, 1.002) | **<.001** |
| White | 11,249 (51.5%) | 195,849 (56.2%) | NR | NR |
| Unavailable | 2,012 (9.2%) | 92,028 (26.4%) | NR | NR |
| SVI, mean (SD) | 0.306 (0.265) | 0.269 (0.243) | 1.001<sup>d</sup><br>(1.001, 1.002) | **<.001** |

<sup>a</sup> aOR calculated relative to the male patients.
<sup>b</sup> aOR calculated relative to the white patients.
<sup>c</sup> Native American, Alaska native, native Hawaiian or other Pacific Islander, and multi-racial.
<sup>d</sup> aOR calculated per every 0.01 point change in SVI.

**Figure 4.**
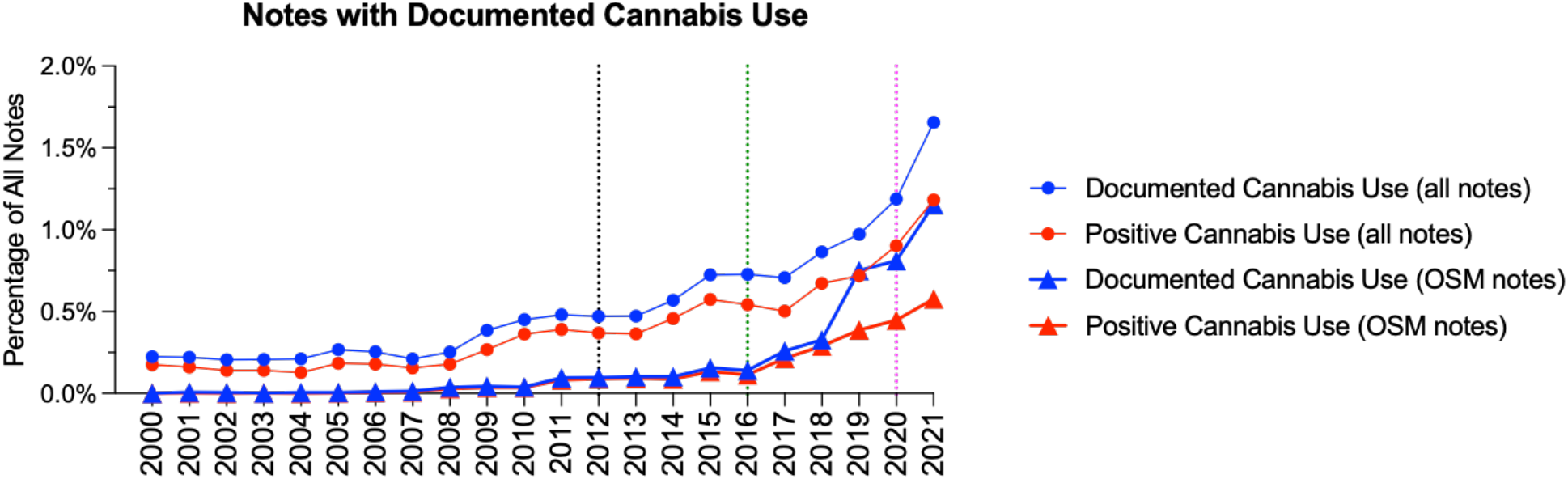
Cannabis use documentation as a percentage of all notes collected each year (circles) and cannabis use documentation in OSM notes as a percentage of all OSM notes collected each year (triangles). The vertical dotted lines indicate the legalization of medical (2012, black) and recreational (2016, green) cannabis in Massachusetts, and declaration of COVID-19 pandemic (2020, magenta).

From 2000 to 2021, there were increases in the total number of new patients with documented positive cannabis use (Figure 5A) with a substantial drop in 2020 corresponding to clinical care restrictions associated with the COVID-19 pandemic (magenta dotted vertical dotted line). Similar trends, with different magnitudes were observed for new patients with documented positive cannabis use within each race category (Figure 5B). While there were no consistent trends in the percentage of new female patients with documented positive cannabis use prior to 2012 (legalization of medical cannabis), the percentage of new female patients with documented positive cannabis use increased consistently from 2012-2021 (Figure 5C). Finally, the average age at first documented cannabis use increased from 2000-2021 (Figure 5D).

**Figure 5.**
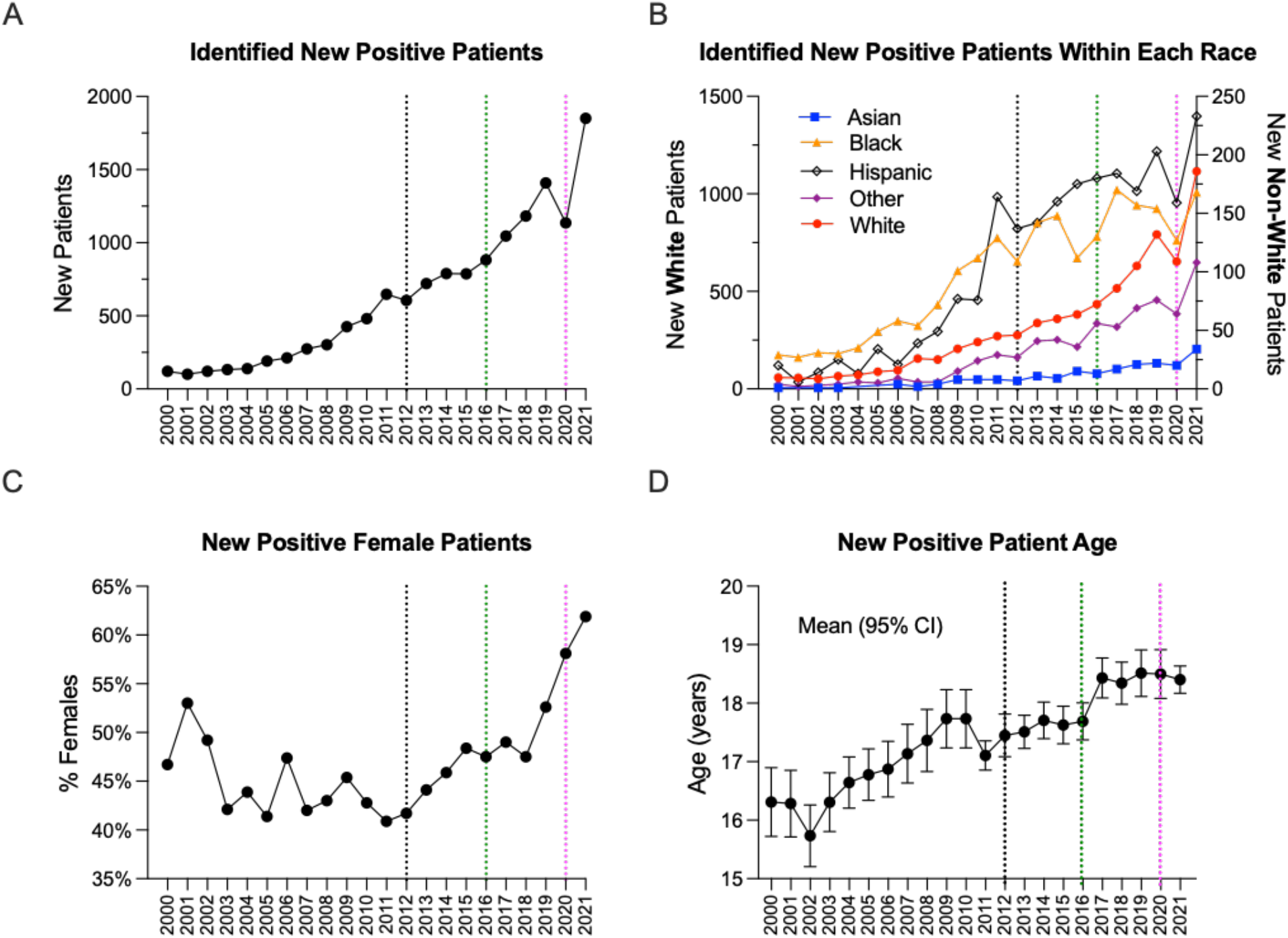
(A) Number of new identified cannabis positive patients per year. (B) Number of new identified cannabis positive patients by each race (other includes Native American, Alaska native, native Hawaiian or other Pacific Islander, and multi-racial). Due to a substantial imbalance in number of white and non-white patients, white patients are plotted on left y-axis and non-white patients are plotted on the right y-axis. (C) New identified female cannabis positive patients as a percentage of all new cannabis positive patients. (D) Average (95% CI) age of new identified cannabis positive patients. The vertical dotted lines indicate the legalization of medical (2012, black) and recreational (2016, green) cannabis in Massachusetts, and declaration of COVID-19 pandemic (2020, magenta).

The distribution of musculoskeletal procedures for both positive and negative cohorts were comparable with application of casting and splints for bone fractures as the most prevalent procedure (Figure 6).

**Figure 6.**
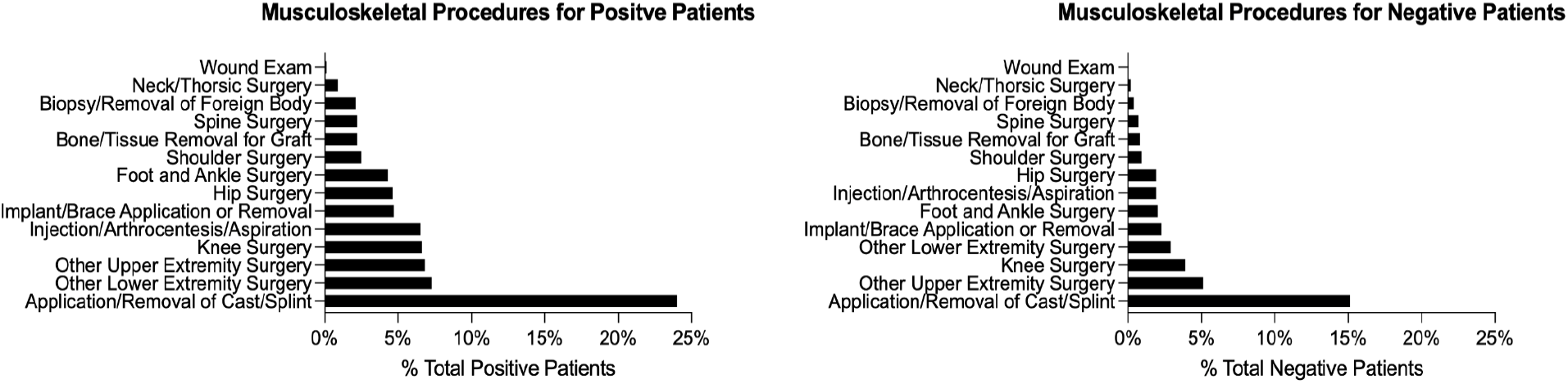
Distribution of common musculoskeletal procedure groups based on CPT billing codes.

Compared to males, females had higher odds of having cannabis use documented in their clinical notes. (aOR=1.074; P<.001; Table 5). Compared to white patients, Asian patients had lower odds (aOR=0.829, P<.001), while black patients (aOR=3.400, P<.001), Hispanic patients (aOR=2.245 P<.001) and those from other racial backgrounds (aOR=1.164, P<.001) had higher odds of cannabis use documentation within clinical notes (Table 5). Patients with higher SVI had higher odds of having cannabis use documented in their clinical notes (aOR=1.001, P<.001; Table 5). We observed similar trends when excluding the medical cannabis use from the patients with documented cannabis use (Supplementary Table S1).

Compared to males, females had lower odds of positive cannabis use (aOR=0.957; P=.038; Table 6). Compared to white patients, Asian patients had lower odds (aOR=0.651, P<.001), while black (aOR=3.222, P<.001) and Hispanic (aOR=2.131, P<.001) patients had higher odds of positive cannabis use (Table 6). Higher SVI was associated with greater odds of positive cannabis use (aOR=1.002, P<.001; Table 6).

**Table 6.** Characteristics of positive cannabis use patients compared to negative patients.

| Characteristics | Positive Patients | Negative Patients | aOR (95% CI) | P-Value |
| --- | --- | --- | --- | --- |
| Total Sample, No. | 13,556 | 356,531 | NR | NR |
| Age at the first documented use, mean (SD), y | 17.9 (5.4) | NR | NR | NR |
| With diagnosed cannabis abuse disorder <sup>a</sup>, No (%) | 1,971 (14.5%) | NR | NR | NR |
| Sex, No (%) |  |  |  |  |
| Female | 6,728 (49.6%) | 188,054 (53.7%) | 0.957 <sup>b</sup><br>(0.918, 0.998) | **.038** |
| Male | 6,827 (50.4%) | 168,440 (47.2%) | NR | NR |
| Unknown | 1 (<0.1%) | 37 (<0.1%) | NR | NR |
| Race, No. (%) |  |  |  |  |
| Asian | 206 (1.5%) | 8,255 (2.3%) | 0.651 <sup>c</sup><br>(0.557, 0.761) | **<.001** |
| Black | 2,144 (15.8%) | 15,618 (4.4%) | 3.222 <sup>c</sup><br>(3.014, 3.444) | **<.001** |
| Hispanic | 2,280 (16.8%) | 22,486 (6.3%) | 2.131 <sup>c</sup><br>(1.994, 2.278) | **<.001** |
| Other <sup>d</sup> | 685 (5.1%) | 17,275 (4.8%) | 1.007 <sup>c</sup><br>(0.918, 1.104) | .888 |
| White | 7,004 (51.7%) | 200,094 (56.1%) | NR | NR |
| Unavailable | 1,237 (9.1%) | 92,803 (26.0%) | NR | NR |
| SVI, mean (SD) | 0.309 (0.266) | 0.269 (0.243) | 1.002 <sup>e</sup><br>(1.001, 1.003) | **<.001** |

<sup>a</sup> Determined based on diagnostic ICD codes.
<sup>b</sup> aOR calculated relative to the male patients.
<sup>c</sup> aOR calculated relative to the white patients.
<sup>d</sup> Native American, Alaska native, native Hawaiian or other Pacific Islander, and multi-racial.
<sup>e</sup> aOR calculated per every 0.01 point change in SVI.

## Discussion

This study demonstrates the utility of NLP in evaluating cannabis use and its documentation from heterogeneous and unstructured clinical notes. The generated large-scale database enabled us to investigate the changes in cannabis use documentation and positivity rate, as well as potential disparities in cannabis use and documentation among children and young adults over a period of 21 years. We observed an increasing trend in cannabis use documentation, in particular over the past decade, with smaller proportions within the OSM clinical notes. These increases included both overall cannabis use documentation (e.g., positive or negative) and documented positive use. Multiple factors may have contributed to these increasing trends. In particular, the legalization of medical and recreational cannabis use, the development of new cannabis-based medications (e.g. Epidiolex, Marinol) as well as the cultural and policy changes may all have contributed to observed increases in cannabis use documentation. Cannabis was legalized for medical use in the state of Massachusetts in late 2012 and for recreational use later in 2016. These legalizations have possibly resulted in an increased cannabis usage [26, 27] and/or greater attention paid to the documentation of cannabis use during visits [28, 29]. Despite increasing trends, a very small portion of notes overall documented cannabis use (<2%), which for the most part contained insufficient information (i.e., duration, frequencies, amount). These findings are in line with recent studies indicating discrepancies between patient reported cannabis (e.g., surveys) and cannabis use documentation in health records [30], while showing a much lower rate than those previously reported.

We saw significant disparities in cannabis use and its documentation. Female patients, non-white non-asian racial groups and those from a more vulnerable socioeconomic backgrounds (i.e., higher SVI) had higher rates of cannabis documentation (whether the patient was asked about cannabis usage or cannabis was discussed with the patient in any way) in their clinical notes. With regards to cannabis use (i.e., positive vs negative), we saw higher rates of cannabis use among male patients, black or Hispanic patients, and those from a more vulnerable socioeconomic background. While the observed discrepancies in use are in agreement with those reported in prior studies [31, 32], the demographic and socioeconomic differences in cannabis documentation highlight potential biases and implementation challenges in proper tracking of cannabis use among children and young adults. The observed racial and socioeconomic disparities in cannabis use documentation may also be an underlying factor for the observed higher positive cannabis use rates among those patients. This highlights the need for a systematic and comprehensive approach to discuss and document cannabis use, regardless of patient demographic and socioeconomic background, which may in turn lead to a more accurate assessment of true disparities in cannabis use.

We saw a comparable distribution of musculoskeletal procedures between cannabis users and non-users. This may suggest that the use of cannabis is not primarily influenced by underlying injuries and health conditions. However, whether the use of cannabis has downstream effects on the treatment choices and outcomes requires further evaluations. The high prevalence of cast/splint procedures, often done for treatment of bone fractures, among cannabis users along with prior reports of changes in bone remodeling due to cannabis use [3, 4, 30, 33–37] further highlight the importance of proper cannabis use documentation to improve treatment outcomes.

There are several limitations which should be considered when interpreting the current findings. The primary limitation of this study is limited generalizability. This study focused on a single institution, in a single state. Other states may have different policies regulating cannabis access, and patients in those states may have different cultural norms around cannabis use and disclosure.

In addition, this study analyzed cannabis usage in a binary (positive/negative) manner since in most cases information on duration, frequency, and dosage were missing. Also, with the exception of toxicology reports and medical cannabis use (e.g., prescription drugs), cannabis use was self-reported which may have biased our observations. Despite these limitations, this study is among the most comprehensive efforts to evaluate cannabis use documentation in a large corpus of clinical notes. Further, to our knowledge, our developed NLP pipeline has the highest performance metrics in capturing a diverse set of cannabis use patterns from clinical notes, which adds to the reliability of the observed findings.

## Conclusions

In this study a hierarchical NLP pipeline was developed and used to conduct an in-depth analysis of cannabis use documentation in a large and heterogeneous corpus of clinical notes obtained over a period of 21 years. The promising performance of the developed NLP pipeline (i.e. AUROC=0.94) supports the use of these approaches in large-scale screening of clinical notes for even complex tasks such as medical and recreational drug use. The findings also highlight the need for rigorous guidelines for proper screening and documentation of cannabis use in electronic health records. Such efforts would enable evidence-based research into the clinical impact of cannabis and support efforts to address observed disparities not only among cannabis users but also in whether patients were asked about cannabis use during their visits.

## Data Availability

The data used in this study cannot be publicly shared due to privacy concerns. Data can be access upon reasonable request pending approvals from Boston Children's Hospital.

## Author Contributions Statement

Study concept and design was done by AMK and GDH. Data collection was done by all authors. Analysis was done by NT and AMK. Interpretation of the findings was done by all authors. First draft of the manuscript was developed by NT, MR, MS, and AMK. All co-authors contributed to the reviewing and finalizing the manuscript.

## Ethical Approval

This study was performed in line with the principles of the Declaration of Helsinki. Approval was granted by the Institutional Review Board of Boston Children’s Hospital.

## Consent to Participate

This a retrospective study and consent was not required as approved by the IRB.

## Supplementary Materials

**Supplementary Table S1.** Characteristics of patients with and without cannabis use documentation excluding medical cannabis use.

| Characteristics | Documented Patients | Not Documented Patients | aOR (95% CI) | P-Value |
| --- | --- | --- | --- | --- |
| Total Sample, No. | 20,312 | 348,248 | NR | NR |
| Sex, No (%) |  |  |  |  |
| Female | 10,739 (52.9%) | 183,311(52.6%) | 1.098 <sup>a</sup><br>(1.061, 1.137) | **<.001** |
| Male | 9,572 (47.1%) | 164,901 (47.4%) | NR | NR |
| Unknown | 1 (<0.1%) | 36 (<0.1%) | NR | NR |
| Race, No. (%) |  |  |  |  |
| Asian | 365 (1.8%) | 8,057 (2.3%) | 0.826 <sup>b</sup><br>(0.735, 0.928) | **.001** |
| Black | 3,303 (16.3%) | 14,403 (4.1%) | 3.713 <sup>b</sup><br>(3.514, 3.924) | **<.001** |
| Hispanic | 3,427 (16.9%) | 21,188 (6.1%) | 2.362 <sup>b</sup><br>(2.236, 2.495) | **<.001** |
| Other <sup>c</sup> | 1,134 (5.6%) | 16,723 (4.8%) | 1.200 <sup>b</sup><br>(1.117, 1.290) | **<.001** |
| White | 10,133 (49.9%) | 195,849 (56.2%) | NR | NR |
| Unavailable | 1,950 (9.6%) | 92,028 (26.4%) | NR | NR |
| SVI, mean (SD) | 0.308 (0.267) | 0.269 (0.243) | 1.001 <sup>d</sup><br>(1.001, 1.002) | **<.001** |

<sup>a</sup> aOR calculated relative to the male patients.
<sup>b</sup> aOR calculated relative to the white patients.
<sup>c</sup> Native American, Alaska native, native Hawaiian or other Pacific Islander, and multi-racial.
<sup>d</sup> aOR calculated per every 0.01 point change in SVI.

